# Balance in Dance: A Scoping Review

**DOI:** 10.1101/2025.06.04.25328967

**Authors:** Miko Gianola Fratnik, Isobel Jupp, Phaedra Petsilas, Peter Dunleavy, Matthew Banger, Alison McGregor, Richard L. Abel

## Abstract

Balance is a fundamental component of dance performance, yet its definition and assessment in dance remain challenging. This scoping review aims to bridge the gap between existing balance research and practical applications by critically evaluating current measurement technologies, examining factors influencing balance performance, and exploring the mechanisms through which these factors interact with dance execution. A comprehensive search of PubMed and Embase databases identified 69 relevant papers published since 1999, focusing exclusively on studies on people who regularly engage in dance. Our analysis revealed limitations in standard balance assessment tools, with *most* current tests failing to sufficiently challenge skilled dancers. Recent developments in dance-specific assessments, particularly modifications of the Star Excursion Balance Test, show promise, though their effectiveness varies across different dance styles and skill levels. Force plate analysis combined with skill-based tests provide a more comprehensive assessment of capabilities, especially with respect to motor skill acquisition and injury recovery. Literature analysis identified several factors affecting balance in dance: aerobic capacity, muscle fatigue, lower extremity strength, and previous injuries. Additionally, technical considerations, such as pointe shoe degradation, significantly impact balance strategies and injury risk. Although strength training shows positive effects on balance and dance-specific skills, the influence of cognitive load, nutrition, and long-term training adaptation remains unclear. Future research should prioritize developing and validating dance-specific balance measures while investigating targeted interventions for different dance populations. This synthesis emphasizes the critical need for dance-specific approaches to balance assessment and training, with direct applications for both research methodology and dance education practices.

## Introduction

Sustained single-leg balance is fundamental to many dance styles, making balance control a critical skill for dancers. Over the past decade, researchers investigated how balance capabilities influence dance performance, as stability is essential for achieving aesthetic movement qualities, particularly in anatomically challenging positions [1]. Professional dancers dedicate substantial training time to developing and maintaining balance skills that exceed performance requirements. However, dancers experience fluctuations in balance abilities, with the underlying causes of these variations remaining poorly understood. Inconsistencies in balance control not only affect performance quality and cause frustration but may also increase injury risk [2,3].

Recent research has advanced our understanding of balance measurement and assessment protocols, yet there remains a critical need to synthesise and translate these findings for both researchers and practitioners [2-4]. Understanding how various factors— including proprioception, aerobic capacity, muscle fatigue, strength, previous injuries, and nutrition—affect balance in dance is crucial for optimising training and performance [4–6]. A scoping review approach is ideal for this purpose, as it enables a comprehensive synthesis of diverse information sources whilst bridging the gap between research and practice. Unlike systematic reviews, which examine specific research questions through standardised methodologies, scoping reviews offer broader perspectives by integrating findings across multiple disciplines. This integration of industry, medical, and academic knowledge provides a holistic understanding of dancers’ balance and highlights areas requiring further investigation.

### Aim and objectives

This scoping review aims to bridge the gap between existing balance knowledge and practical applications. The review critically evaluates current technologies and tests for measuring balance in dance, evaluating the effectiveness and applicability of existing measurement techniques. It examines factors influencing balance performance, including physiological and biomechanical aspects, and explores the mechanisms through which these factors interact with balance and dance performance. Additionally, it aims to define balance in the context of dance, expanding on the generic definition to capture the dynamic and complex nature of dance movements across various styles. Finally, the review identifies future research directions for improved measurement tools and interventions. This includes suggesting new methodologies and technologies that could enhance the accuracy and reliability of balance assessments, refining existing tools, and recommending targeted interventions to support dancers’ balance and overall performance. By addressing these objectives, the review seeks to contribute to the advancement of dance science and practice, ultimately enhancing the health and performance of dancers.

## Methods

A comprehensive search of PubMed and Embase was conducted in June 2024 to identify studies discussing factors influencing balance in dance. The search strategy incorporated specific keywords and Boolean operators to ensure precision and relevance. The keywords included ‘balance’, ‘dance or dancer’, and combinations such as ‘aerobic capacity’ or ‘muscle strength’ or ‘muscle endurance’ or ‘nutrition’ or ‘previous injuries’ or ‘cognitive load’ or ‘self-efficacy’. The search was refined to exclude articles focusing on unrelated populations and those who do not engage in academic dance regularly. This approach yielded 69 initial studies relevant to the topic. Articles were selected based on their pertinence, with no restrictions on study design, and only publications dated after 1999 were included. Eligible studies were reviewed and synthesized scopingly to provide a comprehensive overview of the factors influencing balance in dance, highlighting critical elements such as aerobic capacity, muscle strength, endurance, nutritional aspects and prior injuries. This synthesis aims to present an inclusive and detailed understanding of the subject. In this review, ‘dancers’ refers to individuals aiming for or engaged in a professional dance career. Those dancing recreationally are labelled as ‘recreational dancers,’ while children are categorised by age. This distinction is crucial as the term ‘dancer’ varies in research, affecting its applicability due to distinct characteristics at each level of experience and dance style.

### Balance

#### Definition of balance

Balance is a prerequisite for obtaining the aesthetic parameters necessary of most dance styles. To our knowledge, there is no consensus on a definition of balance in dance that encapsulates the dynamic nature of this skill [1]. Static balance is defined as the ability to maintain the centre of pressure within the limits of the base of support with minimal movement [7] while dynamic balance is defined as the ability to maintain stability during weight shifting via changes in the base of support [8].

This paper proposes to expand on these generic definitions to establish a common understanding of the terminology for dance. These definitions must account for the fact that dance movement is not static but often transitions through different levels, travels through space, and can interact with living and inanimate partners. Static balance in dance could be defined as the ability to maintain the resulting forces on the body within the desired base of support. This would expand the current definition to body positions that, unlike quiet standing, require significant muscle activation to keep the centre of mass within the base of support. Dynamic balance would also integrate the element of weight shifting. Balance is usually acquired during development; it’s a trainable skill through practice and tends to decay with advanced age. Dancers have a greater ability to maintain static balance compared to the general population, but it is unclear how and when this ability is developed through dance training [9].

#### Current Methods for Measuring Balance in Dancers

Drawing from sport science, efforts to capture balance can be broadly divided into two complementary categories: technology and tests. Technology, both wearable and stationary, can be used to digitally measure balance, while tests are the tasks and procedures employed to evaluate balance [10]. Balance has been widely tested in the general population [11]. In the context of dance research, despite recent publications laying the groundwork by improving previous methods [12], the optimal tools for measuring balance are still undetermined.

Balance tests have been identified by researchers working on measuring dancers’ balance by adapting pre-existing tests or creating new ones. Clarke [2021] compared different balance tests—including the original Star Execution Balance Test (oSEBT), Romberg test, Pirouette tests and Airplane test - and concluded, through correlation analysis of the agreement of the test’s scores, that these existing tools were not relevant or sufficiently challenging for dancers [13]. The study used descriptive statistics to underline the need for more thorough research on balance in dance and highlighted the need for a greater sample size to adequately power inferential studies going forward.

Batson (2010) started by modifying the oSEBT [14] with timed tests, cognitive tasks, and sensory changes, but found that these adjustments did not adequately capture university-level dancers’ balance strategies [15]. Building on these earlier studies, Beckman, since 2022 has made notable progress in adapting the oSEBT for dancers. In 2022, Beckman introduced a dance-specific version of the oSEBT (dsSEBT), incorporating ballet and contemporary dance movements by limiting trunk rotation and adjusting foot positions to improve specificity [16]. While these changes did not significantly affect reach distances, they lead to higher error rates, recorded through a proposed scoring system. This suggested the test had become more challenging and better aligned with dancers’ needs by emphasising proprioceptive feedback.

In 2023, Beckman explored how tempo and the order of movements affected the dsSEBT [17]. The study found that varying the tempo (beats per minute) did not significantly impact reach distances, centre of pressure, or error scores, indicating that tempo alone isn’t crucial for assessing dancers’ balance. However, the study emphasised the importance of completing the full test sequence, as opposed to simplifying it through isolating specific directions as in the Y-Balance Test (YBT), underscoring the need for a comprehensive approach. By 2024, Beckman synthesised these insights to create a practical and challenging balance test for dancers, incorporating the most effective modifications from previous research [12]. This latest version introduced distinct difficulty levels and found a strong correlation between performance on the oSEBT and the new dance-specific stages of the dsSEBT. Despite the study’s small sample size and its focus on ballet and contemporary dance university students, it suggests that further validation and adaptations to different dance styles are needed. Beckman’s work marks a significant advance in developing a more accurate and challenging balance assessment for dancers.

The ‘Airplane test’ is a clinical test used to assess single-leg balance, and is a more advanced variant of the single leg step down test [13]. In this test, the trunk is leaned forward, and the non-support leg is extended backwards, with the pelvis kept parallel to the ground. The participant performs five controlled pliés (knee flexion parallel to the foot) while horizontally adducting the arms to touch the fingertips to the ground [13,18]. The Airplane test assesses whether neutral lower extremity alignment is maintained in at least four out of five pliés. A key advantage is that it requires no additional equipment and is quick to administer.

The Airplane test has been proposed as a possible indicator of a dancer’s functional balance and skill level [13, 19, 20], however the test has been shown to not be a reliable indicator. Richardson e*t. al*. [2010] compared, among other things, the Airplane test against teacher assessment of pointe shoe readiness and did not find agreement on young vocational dance students’ [12.3 ± 2.2 years] pointe shoe readiness between the two. There was some agreement on the lack of readiness among vocational students under twelve. This suggests that passing the Airplane test may be necessary but not sufficient for teachers to assess pointe shoe readiness. The main limitation is its moderate difficulty, making it less suitable for detecting small changes. It could serve as a clinical cut-off, but its applicability requires further research.

#### Technologies to assess balance

To improve the accuracy of the balance measures explored above, it is possible to integrate these tests with technologies, which enable a greater level of detail in assessment. Currently, one of the most widely used technologies to assess balance in dance is force plates. This technology measures Ground Reaction Forces (GRF): the forces created against the standing surface during static and dynamic movement. Through GRF measurements of the GRF it is possible to obtain the centre of pressure (COP), which is the projection on the floor of the sum of forces at the point where the total forces are applied. The COP is used to measure postural control and balance.

Researchers have used COP during turns to describe the difference in balance strategies between novice (12.0 ± 1.9 years) and experienced (17.8 ± 3.4 years) female adolescent ballet students, determining that older students were better able to control the trajectory of the COP [21]. Additional research investigated how feedback affects pirouette (Table 4) performance [16]. Using COP, no difference was found between giving or not giving feedback on the quality of pirouette execution. Nevertheless, participants believed that having feedback [internal or external depending on the individual performer] on their executions positively impacted their performances. This shows that dancers’ perception of balance and performance is subjective, and force plates can provide them with objective measurements.

**Table 1.**
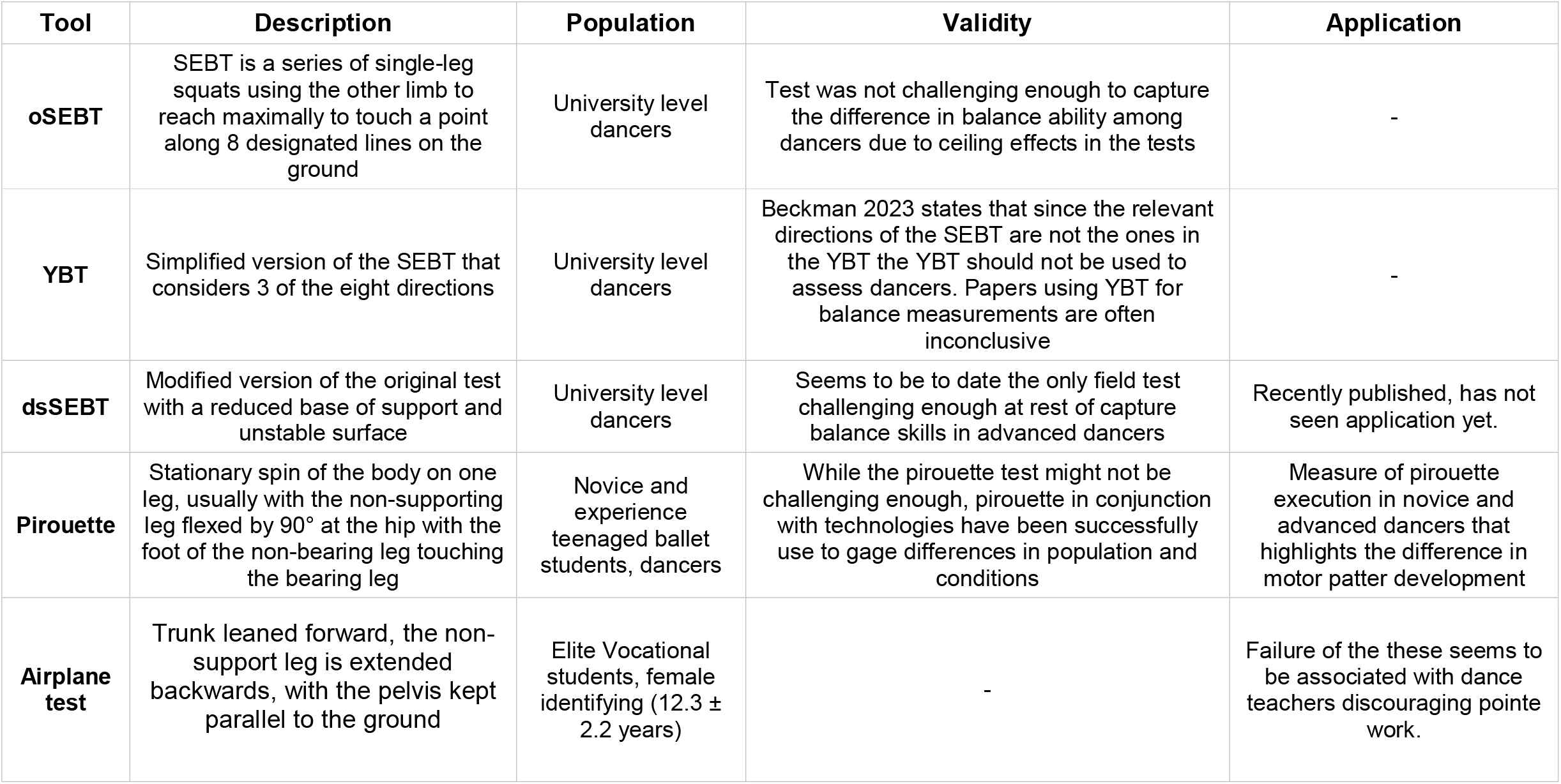
Factors impacting balance in dance

**Table 2.** Technologies measuring balance in dance

| **Technology** | **Parameter** | **Tool Used** | **Population** | **Findings** |
| --- | --- | --- | --- | --- |
| **Force plates** | COP | COP used to measure difference in pirouette execution | Novice and experiences teen aged ballet students, dancers | While the pirouette test might not be challenging enough, pirouette in conjunction with technologies have been successfully used to gauge differences in population and conditions. |
|  | COP sway | See the effect of fatigue protocol on COP during relevé | Advance dance student and professional in ballet and jazz or contemporary dance | Significant differences were found in COP sway during single leg stance in passè following HIDPFT |
| **EMG** | Muscle activation patterns | Determining effect of pointe shoe deterioration | Professional female identifying ballet dancers | A higher tibialis anterior activation was found during arabesque while wearing 'dead' pointe shoes vs new shoes. |
| **Optoelectronics** | Joints angle | Infrared cameras and reflective markers | Teenaged female identifying ballet student | Differences in hip and trunk kinematic in relevé were observed after fatigue protocol |

**Table 3.** Factors impacting balance in dance

| **Factors** | **Association** | **Proposed Mechanism** | **Dance Implication** |
| --- | --- | --- | --- |
| **Aerobic Fatigue** | Aerobic capacity influences balance in professional dances increasing COP displacement. | Increased respiratory demands, among other things influence balance stability | Deliberately incorporating balance training under high aerobic demands could improve balance performance when aerobically fatigued |
| **Muscle Fatigue** | found that fatigue brought ballet dancer to rely more on proximal joint to execute relevé compared to pre-fatigue, resulting in a greater COP displacement. | Muscle necessities for the management of ankle reaction to maintain balance are less efficient due to neuromuscular fatigue happening at the neuromuscular junction | Training muscle fatigue, when roles demand for it, could be an effective way to increase balance. |
| **Strength** | Lower limb and core strength training might improve dance performance. | Musculoskeletal adaptation and improved trunk stabilization. | Incorporating strength training in dance program might result in better dance performance |
| **Previous injuries** | Previous ankle sprains and injuries have a significant relationship with balance | Ankle sprains impede the entire lower chain, and any ligament damage would alter proprioceptive abilities and strength | Exercise intervention combined with blood flow restriction improved ankle stability and functionality, as measured by the Foot and Ankle Ability Measure |
| **Nutrition** | Observing at artistic gymnastics it was found that Carbohydrate solution was able to supply muscle demands and improve the athlete's focus showed by the reduced number of falls when under fatigue | Without supplementation most of the glucose is consumed by the CNS during exercise that is both cognitively and physically demanding, making it less available for muscle use. Supplementing would allow for enough carbohydrates for both functions. | Adequate carbohydrate supply during training and performance could improve performance among dancers. Timing and specificity of nutrition needs to be explored for different dance styles, populations and skill levels. |

**Table 4.**
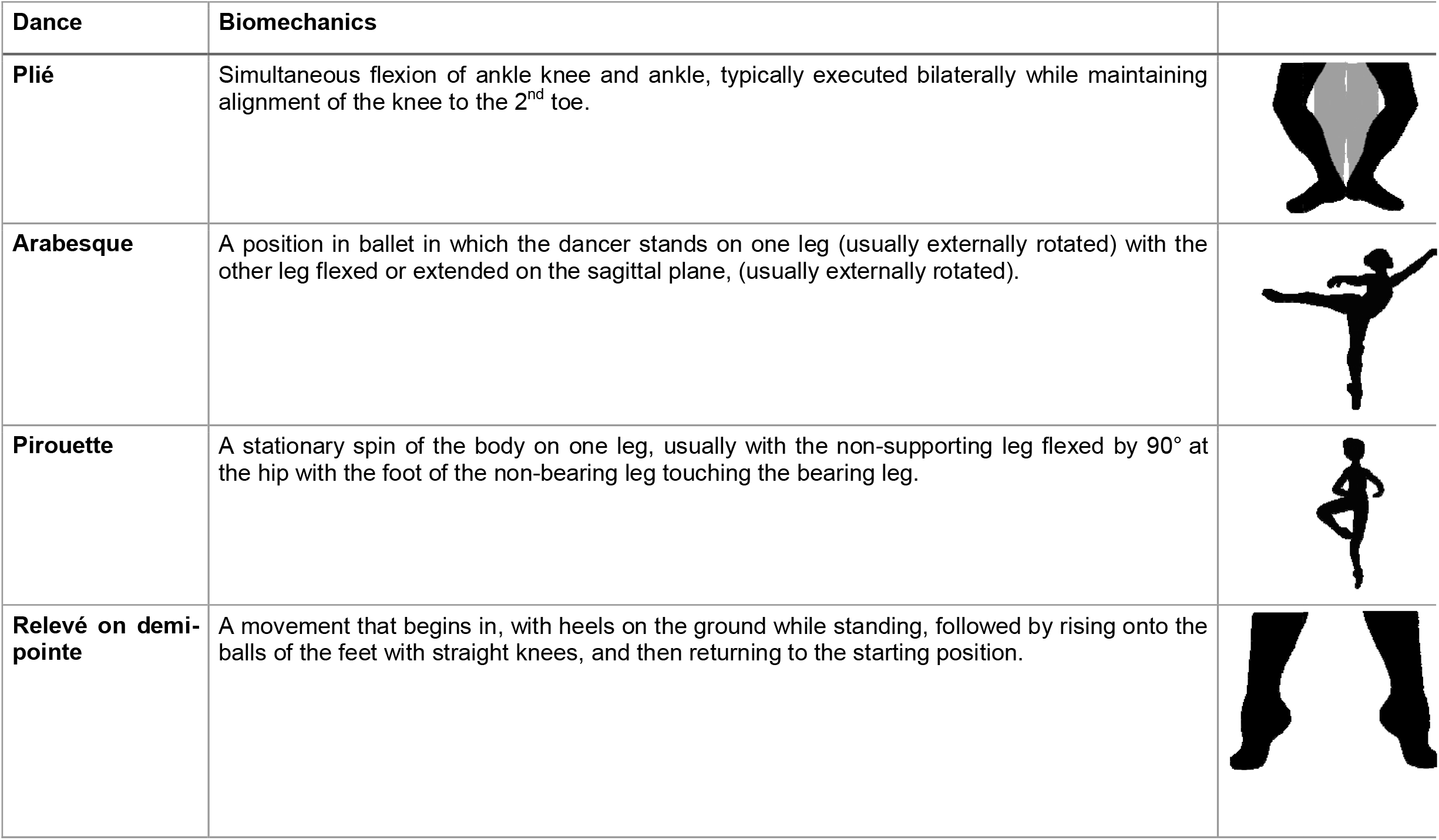
Dance terminology and positions

| Dance | Biomechanics |
| --- | --- |
| **Plié** | Simultaneous flexion of ankle knee and ankle, typically executed bilaterally while maintaining alignment of the knee to the 2<sup>nd</sup> toe. |
| **Arabesque** | A position in ballet in which the dancer stands on one leg (usually externally rotated) with the other leg flexed or extended on the sagittal plane, (usually externally rotated). |
| **Pirouette** | A stationary spin of the body on one leg, usually with the non-supporting leg flexed by 90° at the hip with the foot of the non-bearing leg touching the bearing leg. |
| **Relevé on demi-pointe** | A movement that begins in, with heels on the ground while standing, followed by rising onto the balls of the feet with straight knees, and then returning to the starting position. |

Other researchers have used force plates together with Electromyography (EMG) and optoelectronic systems to document increases of COP sway after fatigue protocol [22]. Similar procedures were carried out to examine differences in balance strategies when comparing new and dead pointe shoes [23]. Dead pointe shoes are shoes that have been worn and no longer offer support for the foot while en pointe. This study compared lower extremity biomechanics, including sway, muscle recruitment, and ankle stability, in professional ballet dancers performing relevé and arabesque in both new and dead pointe shoes (Table 4). The aim was to assess whether shoe deterioration impacted biomechanics, similar to how worn-out running shoes affect runners. Increased sway and tibialis anterior activation in the results could suggest that worn pointe shoes compromise stability as dancers compensate for the diminished support resulting in an increased risk for injury. Identifying degradation thresholds is essential for understanding their impact on biomechanics and balance.

#### Factors Influencing Balance Performance and the underlying mechanisms in dance

Dance technique is not the only factor which determines balance, there are many demands affecting a dancer’s ability to balance. When performing, dancers must be aware of their surroundings and environment, including tracking the music, other performers, and stage staff, whilst simultaneously preparing for the upcoming choreography.

Aerobic capacity and cardiovascular fitness may impact balance in dance. Among the general population, research has shown that that after short-term intensive exercises, hyperventilation, rather than fatigue, is responsible for increased postural sway [24]. Further, studies found that balance was also disrupted after general post-exercise aerobic fatigue [25]. However, the authors reported no difference before and after fatigue in the YBT using the *Dance Aerobic* Fitness *Test* (*DAFT*) [26] protocol. This might be due to two reasons: the DAFT might not effectively exhaust dances’ aerobic systems, and the YBT might not effectively measure changes in balance and postural control due to ceiling effects. Supporting this, Bulkey found that by using the high-intensity dance performance fitness test (HIDPFT) enabled detection of changes in COP in professional dancers (ballet, jazz, or contemporary) [22]. Understanding how different responses to exercise individually contribute to balance acquisition and adaptation would help minimise their effect on dancers.

To date, no study has been able to measure the effects of cognitive load and mental fatigue on dance performance. Researchers found that standard cognitive load protocols such as the Stroop tests were not taxing enough to influence balance in young healthy people [6,27]. Similarly, Baston (2010) [15] tried to integrate a cognitive load component into the dsSEBT and found that the variability in responses did not allow for accurate measurement of the SEBT, nor was it possible to deduce any conclusion on the impact of cognitive load on balance performance.

Strength has recently been investigated in dance research in the context of jumping in elite ballet dancers [28], but its relation with balance in dance remains debated. One study [29] examined whether core and lower limb strength training could improve balance and dance-specific skills, like pirouettes, in university-level dancers using a program based on Jeffreys’ core stabilisation techniques [30]. After nine weeks, dancers showed improved oSEBT, anti-rotation abilities, and pirouette performance. However, it was unclear whether these gains were from dance classes alone or were the result of added strength training. Additional trunk stabilisation work may play a role in improving both static and dynamic balance performance as well as dance-specific performance. Future studies could measure the effect of similar strength training protocols on other styles of dance trough case-control study to isolate its impact, refining strength and conditioning programs in different dance populations.

Some research has investigated the association between strength and aesthetic competence and the relationship between aerobic conditioning and dance performance but the specific relationship between those and balance is yet to be understood [31,32].

Strength and muscle fatigue are different measures of a muscle’s ability to generate force. Research by Lin *et al*. (2016) [33] explored how lower leg muscle fatigue affects balance and biomechanics in dancers during movements like relevé. The research was motivated by the belief that muscle fatigue impairs sensory feedback, affecting balance and increasing injury risk. The findings showed that lower limb fatigue altered biomechanics, particularly in the medial-lateral axis, and dancers shifted body weight to compensate. The study suggests that targeted conditioning programs could train muscle endurance to mitigate these biomechanical changes and reduce fatigue-related injuries.

Muscle fatigue appears to affect balance and muscle activation patterns, especially during repeated weight-bearing plantar flexion exercises. Differences in muscle metabolism may explain these findings. The soleus, rich in slow-twitch oxidative fibres, resists fatigue better than the gastrocnemius, [33] supporting its key role in sustaining relevé. While the gastrocnemius acts as the prime mover, future research could examine fatigue effects on core stabilizers, hip flexors, and external rotators, which contribute to dancer stability.

Injuries are common in dance, with injuries in elite ballet dancers having an incidence rate of 3.9 (per 1000 hours) making it similar to cricket but lower than rugby union or ice hockey [34], with foot and ankle injury accounting for up to 62% of all injuries [35 - 40]. To achieve positions like relevé and arabesque on pointe, ballet and ballet-derived styles require a greater ankle mobility range than the general population [41]. This could be a contributing factor to the high prevalence of foot and ankle injuries among ballet dancers which, long term, can lead to CAI and could hinder balance due to altered muscle activation patterns in proximal joints [33].

Hung *et al*. (2021) investigated the prevalence of ankle instability in dancers (ballet) and its relationship with dynamic balance and lower extremity strength [42]. The researchers aimed to explore whether ankle instability correlated with decreased motor proficiency. The results indicated that dancers fell between the proposed cut-offs for chronic ankle instability (CAI) but showed no significant interaction in single-leg balance control and leg dominance. Contrary to expectations, the study inferred that ankle instability did not impact strength. The study may have been limited by the lack of challenging positions for dancers. Furthermore, Hung’s findings did not agree with extensive research on both the general population [43] and high-performing athletes that finds CAI affects the lower extremity muscle chain [44]. The discrepancy may be due to the sensitivity of the measurement tools used in the study. Therefore, It is crucial to study dancers using advanced technologies to further explore the link between balance, prior ankle injuries, and CAI. There is potential to benefit from strength training for ankle injuries in dancers, as research [45] demonstrated that strength exercise intervention combined with blood flow restriction improved ankle stability and functionality, as measured by the Foot and Ankle Ability Measure (FAAM) [46]. This indicates that strength training, which likely enhances balance, leads to better functional outcomes in dancers with chronic ankle instability.

Although there is literature available on dancer’s nutritional intake, little has been published on the effect of nutrition on physical performance in dance. Even when expanding research to other aesthetic sports, is evident that further research is necessary. Research in artistic gymnastics found that carbohydrate supplementation was able to supply muscle demands and improve athlete’s focus, as shown by the reduced number of falls when under fatigue [6,47,48]. Similar studies should be replicated in different dance situations, such as full-length ballet performances or other styles that require repeated intermittent efforts and significant mastery of balance, to determine if similar findings apply to dance.

### Proposed areas for practical application of future research and in dance education

Among the objectives for this review was proposing areas for future research to develop better measurement tools and interventions. Building on previous studies by Beckman (2024), a larger sample size is needed to further validate the SEBT and test the efficacy of the step approach taken in stratifying the 2024 version of the test [12]. Furthermore, since the test has been developed specifically for ballet and contemporary university dance students, validation and adaptations are needed for other populations. Future research should strengthen this new test by increasing the sample size and adapting it to other dance styles. Beckman’s work marks a significant advance in developing a more accurate and challenging balance assessment. As some universities offering dance programs implement screenings across their student population to monitor their well-being and progress, the SEBT and force plates could be incorporated into existing protocols to improve the specificity and reliability of these longitudinal data acquisitions.

Pointe shoes are a significant component of ballet training. The papers examined in this review aimed to validate ballet teachers’ assessment through clinical tests but were unsuccessful. Building on that work, future research could focus on the application of guidelines for pointe shoes readiness together with standardisation of some of the criteria such as functional core strength and ankle mobility [49,50]. Regarding pointe shoes, COP measurements have used to determine changes of biomechanics when wearing new and dead shoes; further research could use similar methods to determine the threshold for ‘dead’ pointe shoes and potentially put systems in place to reduce associated acute and chronic injuries. In several sports, including football, rugby, skiing, and running, technology allows for the mass production of shoes to be built according to athletes’ unique anthropometric parameters, including the insertion of tracking devices in footwear. Given the recent innovations in pointe shoe manufacturing, similar technology could be implemented in dance to help reduce strain on the body and improve performance.

Regarding aerobic demand measurement, the HIDPFT test appears to challenge dancers to the point of compromising balance. Identifying which component is most demanding could help minimize its impact, potentially improving balance and overall performance [22]. Among the general population aerobic capacity impacts balance, with hyperventilation causing increased postural sway after intense exercise, while muscle fatigue shifts balance reliance to proximal joints [24].

Nutrition’s impact on dance performance remains under-explored, but evidence from related sports suggests proper nutrition could enhance dancers’ focus and performance. Researchers could implement an initial case-control study to understand the impact of glucose supplementation during long bouts of activity in dance, such as rehearsals and full-length performances, measuring injury rates and perceived exertion. Subsequent studies could explore how glucose supplementation affects balance specifically or other aspects of fitness that subsequently influence balance, such as aerobic fitness and muscle fatigue. To better translate the research into practical applications, studies should examine the timing, frequency, and concentration of the glucose supplementation. The general guidelines of 30-60g of carbohydrates per hour could be a starting point [51].

Regarding guidelines for dance education, this review suggests that training lower limb muscle fatigue could be an effective way to improve balance, potentially allowing for better overall results. This could be done by running the same routine multiple times in a row or by training a specific muscle group separately from a dance context to improve muscle endurance while dancing. For dance students with ankle sprains exercise intervention combined with blood flow restriction improved ankle stability and functionality, this training could be supplemental to the dance classes or could occur during said classes in case of a recent injury as a replacement for “sitting out” part of classes. Finally, adequate carbohydrate supply during training and performance could improve performance among dancers. Research needs to explore timing and specificity of nutrition requirements for different dance styles, populations, and skill levels.

## Conclusion

This scoping review has highlighted the complexity of balance in dancers, emphasised the need for a more comprehensive and dance-specific understanding of balance performance. The findings underscore the limitations of current balance measurement tools and the importance of developing dance-specific definitions and assessments that can capture the nuanced demands of various dance styles. Factors such as muscle fatigue, strength, previous injuries, and potentially nutrition have been identified as significant influences on dancers’ balance, suggesting that a holistic approach to training and performance preparation is crucial. The integration of advanced technologies, like force plates with genre-specific movements, offers promising avenues for more accurate balance assessment, particularly in monitoring motor skill acquisition and injury recovery. However, the review also reveals significant gaps in our current knowledge, particularly in understanding the effects of cognitive load and the long-term impact of dance training on balance. As the field of dance science continues to evolve, future research should focus on [1] developing more sensitive and specific balance measures for dancers, [2] exploring the underlying mechanisms of balance adaptation in dance training, and [3] investigating targeted interventions that can enhance both performance and injury prevention. By advancing our understanding of balance in dance, we can not only improve the training and performance of dancers but also potentially contribute to broader applications in physical therapy, fall prevention, and general fitness.

## Data Availability

All data produced in the present study are available upon reasonable request to the authors

## Notes

### Competing Interest Statement

The authors have declared no competing interest.

### Funding Statement

This study did not receive any funding

## References

1. Clarke FA. Balance Performance Of Undergraduate Dancers: An Evaluation Of Current And Novel Approaches In Balance Testing And Training In Theatrical Dance. University of Wolverhampton; 2020.

2. Gribble PA, Hertel J, Plisky P. Using the Star Excursion Balance Test to Assess Dynamic Postural-Control Deficits and Outcomes in Lower Extremity Injury: A Literature and Systematic Review. Journal of Athletic Training. 2012 May 1;47(3):339–57.

3. Kenny SJ, Whittaker JL, Emery CA. Risk factors for musculoskeletal injury in preprofessional dancers: a systematic review. Br J Sports Med. 2016 Aug;50(16):997–1003.

4. Svantesson, Ulrika Österber U. MUSCLE FATIGUE IN A STANDING HEEL-RISE TEST. Scandinavian Journal of Rehabilitation Medicine. 1998 Aug 12;30(2):67– 72.

5. Chua TX, Sproule J, Timmons W. Effect of Skilled Dancers’ Focus of Attention on Pirouette Performance. j dance med sci. 2018 Sep 15;22(3):148–59.

6. Batatinha HAP, Da Costa CE, De França E, Dias IR, Ladeira APX, Rodrigues B, et al. Carbohydrate use and reduction in number of balance beam falls: implications for mental and physical fatigue. Journal of the International Society of Sports Nutrition. 2013 Jan 3;10(1):32.

7. Dunsky A, Zeev A, Netz Y. Balance Performance Is Task Specific in Older Adults. BioMed Research International. 2017;2017:1–7.

8. O’Sullivan SB, Schmitz TJ, Fulk GD. Physical rehabilitation. Sixth edition. Philadelphia: F.A. Davis Company; 2014. 1505 p.

9. Golomer E, Dupui P. Spectral Analysis of Adult Dancers’ Sways: Sex and Interaction Vision - Proprioception. International Journal of Neuroscience. 2000 Jan;105(1–4):15–26.

10. Ani KU, Ibikunle PO, Nwosu CC, Ani NC. Are the Current Balance Screening Tests in Dance Medicine Specific Enough for Tracking the Effectiveness of Balance-Related Injury Rehabilitation in Dancers? A Scoping Review. j dance med sci. 2021 Dec 15;25(4):217–30.

11. Promsri A. Analysis of bilateral muscle coordination for characterizing neuromuscular function in postural control. MethodsX. 2022;9:101944.

12. Beckman S, Brouner J. The Development and Reliability of an Updated Dance-Specific Star Excursion Balance Test Protocol. Journal of Dance Medicine & Science. 2024 Jul 28;1089313×241265237.

13. Clarke F, Koutedakis Y, Wilson M, Wyon M. Associations Between Static and Dynamic Field Balance Tests in Assessing Postural Stability of Female Undergraduate Dancers. j dance med sci. 2021 Sep 15;25(3):169–75.

14. Kaminski TW, Gribble P. The Star Excursion Balance Test as a Measurement Tool. Athletic Therapy Today. 2003 Mar;8(2):46–7.

15. Batson G. Validating a Dance-specific Screening Test for Balance: Preliminary Results from Multisite Testing. Medical Problems of Performing Artists. 2010 Sep 1;25(3):110–5.

16. Beckman S, Brouner J. Developing the Positional Characteristics of a Dance-Specific Star Excursion Balance Test (dsSEBT). j dance med sci. 2022 Mar 15;26(1):50–7.

17. Beckman S, Brouner J. Developing the Temporal and Order Characteristics of a Dance-Specific Star Excursion Balance Test (dsSEBT). Journal of Dance Medicine & Science. 2023 Dec;27(4):232–40.

18. Vaganova AI. Basic principles of classical ballet: Russian ballet technique. New York: Dover Publications; 1969. 171 p.

19. Richardson M, Liederbach M, Sandow E. Functional Criteria for Assessing Pointe-Readiness. Journal of Dance Medicine & Science. 2010 Sep;14(3):82–8.

20. DeWolf A, McPherson A, Besong K, Hiller C, Docherty C. Quantitative Measures Utilized in Determining Pointe Readiness in Young Ballet Dancers. j dance med sci. 2018 Dec 1;22(4):209–17.

21. Lin CW, Su FC, Lin CF. Kinematic Analysis of Postural Stability During Ballet Turns (pirouettes) in Experienced and Novice Dancers. Front Bioeng Biotechnol. 2019 Oct 25;7:290.

22. Bulkley D, Jarvis DN. Dancers exhibit decreases in postural control after fatigue. Sports Biomechanics. 2023 May 12;1–12.

23. Aquino J, Amasay T, Sue Shapiro, Kuo YT, Ambegaonkar JP. Lower extremity biomechanics and muscle activity differ between ‘new’ and ‘dead’ pointe shoes in professional ballet dancers. Sports Biomechanics. 2021 May 19;20(4):469–80.

24. Zemková E, Hamar D. Physiological Mechanisms of Post-Exercise Balance Impairment. Sports Med. 2014 Apr;44(4):437–48.

25. Armstrong R, Brogden CM, Milner D, Norris D, Greig M. The Influence of Fatigue on Star Excursion Balance Test Performance in Dancers. j dance med sci. 2018 Sep 15;22(3):142–7.

26. Wyon M, Head A. Development, Reliability, and Validity of a Multistage Dance Specific Aerobic Fitness Test (DAFT). 2003;7(3):5.

27. Salihu AT, Usman JS, Hill KD, Zoghi M, Jaberzadeh S. Mental fatigue does not affect static balance under both single and dual task conditions in young adults. Exp Brain Res. 2023;241(7):1769–84.

28. Mattiussi AM, Shaw JW, Cohen DD, Price P, Brown DD, Tallent J. Reliability, variability, and minimal detectable change of bilateral and unilateral lower extremity isometric force tests. 2022;6(3):9.

29. Watson T, Graning J, McPherson S, Carter E, Edwards J, Melcher I, et al. DANCE, BALANCE AND CORE MUSCLE PERFORMANCE MEASURES ARE IMPROVED FOLLOWING A 9-WEEK CORE STABILIZATION TRAINING PROGRAM AMONG COMPETITIVE COLLEGIATE Dancers. Int J Sports Phys Ther. 2017 Feb;12(1):25–41.

30. Jeffreys I. Developing a Progressive Core Stability Program. National Strength & Conditioning Association. 2002 Oct;24(5):65–6.

31. Koutedakis Y, Hukam H, Metsios G, Nevill A, Giakas G, Jamurtas A, et al. The Effects of Three Months of Aerobic and Strength Training on Selected Performance- and Fitness-Related Parameters in Modern Dance Students. J Strength Cond Res. 2007;21(3):808.

32. Angioi M, Metsios, George, Twitchett E, Koutedakis Y, Wyon M. Association between selected physical fitness parameters and esthetic competence in contemporary dancers. Journal of Dance Medicine & Science. 2009;

33. Lin CF, Lee WC, Chen YA, Hsue BJ. Fatigue-Induced Changes in Movement Pattern and Muscle Activity During Ballet Releve on Demi-Pointe. Journal of Applied Biomechanics. 2016 Aug;32(4):350–8.

34. Mattiussi AM, Shaw JW, Williams S, Price PD, Brown DD, Cohen DD, et al. Injury epidemiology in professional ballet: a five-season prospective study of 1596 medical attention injuries and 543 time-loss injuries. Br J Sports Med. 2021 Aug;55(15):843–50.

35. Garrick JG, Requa RK. An analysis of epidemiology and financial outcome. Am J Sports Med. 1993 Jul;21(4):586–90.

36. Bronner S, Ojofeitimi S, Rose D. Injuries in a Modern Dance Company: Effect of Comprehensive Management on Injury Incidence and Time Loss. Am J Sports Med. 2003 Mar;31(3):365–73.

37. Smith PJ, Gerrie BJ, Varner KE, McCulloch PC, Lintner DM, Harris JD. Incidence and Prevalence of Musculoskeletal Injury in Ballet: A Systematic Review. Orthopaedic Journal of Sports Medicine. 2015 Jul 1;3(7):2325967115592621.

38. Ojofeitimi S, Bronner S. Injuries in a modern dance company effect of comprehensive management on injury incidence and cost. J Dance Med Sci. 2011 Sep;15(3):116–22.

39. Ramkumar PN, Farber J, Arnouk J, Varner KE, Mcculloch PC. Injuries in a Professional Ballet Dance Company: A 10-year Retrospective Study. J Dance Med Sci. 2016 Mar 15;20(1):30–7.

40. Katakura M, Kedgley AE, Shaw JW, Mattiussi AM, Kelly S, Clark R, et al. Epidemiological Characteristics of Foot and Ankle Injuries in 2 Professional Ballet Companies: A 3-Season Cohort Study of 588 Medical Attention Injuries and 255 Time-Loss Injuries. Orthopaedic Journal of Sports Medicine. 2023 Feb 1;11(2):23259671221134131.

41. Khan K, Roberts P, Nattrass C, Bennell K, Mayes S, Way S, et al. Hip and Ankle Range of Motion in Elite Classical Ballet Dancers and Controls. Clinical Journal of Sport Medicine. 1997 Jul;7(3):174.

42. Hung YJ, Boehm J, Reynolds M, Whitehead K, Leland K. Do Single-Leg Balance Control and Lower Extremity Muscle Strength Correlate with Ankle Instability and Leg Injuries in Young Ballet Dancers? j dance med sci. 2021 Jun 15;25(2):110–6.

43. Hall EA, Docherty CL, Simon J, Kingma JJ, Klossner JC. Strength-Training Protocols to Improve Deficits in Participants With Chronic Ankle Instability: A Randomized Controlled Trial. Journal of Athletic Training. 2015 Jan 1;50(1):36– 44.

44. Thompson C, Schabrun S, Romero R, Bialocerkowski A, Van Dieen J, Marshall P. Factors Contributing to Chronic Ankle Instability: A Systematic Review and Meta-Analysis of Systematic Reviews. Sports Med. 2018 Jan;48(1):189–205.

45. Liu Z, Gong Y, Nagamoto H, Okunuki T, Yamaguchi R, Kobayashi Y, et al. Low Body Fat Percentage and Menstrual Cycle Disorders in Female Elite Adolescent Dancers. Journal of Dance Medicine & Science. 2024 Jun;28(2):109–16.

46. Martin RL, Irrgang JJ, Burdett RG, Conti SF, Swearingen JMV. Evidence of Validity for the Foot and Ankle Ability Measure (FAAM). Foot Ankle Int. 2005 Nov;26(11):968–83.

47. Bladt F, Varaeva YR, Benjamin J, Patel S, Courtney A, Holloway P, et al. Preliminary study of bone biomarkers in elite female ballet dancers. CISS. 2023 May 16;8(1):004.

48. Bladt F, Varaeva YR, Retter G, Courtney A, Holloway PAH, Frost G, et al. Pilot study of bone turnover biomarkers, diet, and exercise in elite female ballet dancers. 2024;

49. Weiss DS, Rist RA, Grossman G. Guidelines for Initiating Pointe Training. International Association for Dance Medicine & Science [Internet]. Available from: https://iadms.org/media/5779/iadms-resource-paper-guidelines-for-initiating-pointe-training.pdf

50. hough-coles K, Wyon M. Determining Pointe Readiness in Young Adolescent Female Dancers: A Systematic Review. Journal of Dance Medicine & Science. 2022 Sep 15;26.

51. Jeukendrup A. A Step Towards Personalized Sports Nutrition: Carbohydrate Intake During Exercise. Sports Med. 2014 May;44(S1):25–33.

